# Applying the SEIR Model in Forecasting The COVID-19 Trend in Malaysia: A Preliminary Study

**DOI:** 10.1101/2020.04.14.20065607

**Authors:** Aidalina Mahmud, Poh Ying Lim

**Affiliations:** Department of Community Health, Faculty of Medicine and Health Sciences, Universiti Putra Malaysia, Selangor, Malaysia

**Keywords:** SEIR model, COVID-19, forecasting, trend, Malaysia

## Abstract

On March 18, 2020 the Malaysian government implemented a 14-day Movement Control Order (MCO) as part of the mitigation plan in controlling the COVID-19 epidemic in the country. The MCO aims to limit the contact rates among the population and hence prevent the surge of infected individuals. However, the trend of the epidemic before and after the MCO was not apparent. By applying the Susceptible, Exposed, Infectious and Removed (SEIR) mathematical model, we aimed to forecast the trend of COVID-19 epidemic in Malaysia using data from March 17 to 27, 2020. Based on several predetermined assumptions, the results of the analyses showed that after the implementation of the 14-day MCO from March 18 to 31, 2020, it is forecasted that the epidemic in Malaysia will peak approximately in the end of April 2020 and will subside by about the first week of July 2020. The MCO will “flatten the epidemic curve” but will prolong the duration of the epidemic. Decision to extend the duration of the MCO should depend on the consideration of socioeconomic factors as well.

**Author summary:** Dr. Aidalina Mahmud is a Public Health Specialist and a medical lecturer in the Department of Community Health, Universiti Putra Malaysia. Dr. Lim Poh Ying is a Biostatistician and is a senior lecturer in the Department of Community Health, Universiti Putra Malaysia.

## Introduction

In Malaysia, the COVID-19 pandemic progressed from the alert phase when the nation was made aware of the epidemic in China in late 2019, to containment phase when the disease arrived in Malaysia in mid-January 2020. Malaysia announced its first Covid-19 cases on January 25 involving three China tourists who had come into Malaysia from Singapore on January 23[1]. Up until the first week of February, the positive cases were all epidemiologically linked to someone from China of who has been to China. Two individuals from the humanitarian mission to Hubei were also tested positive for COVID-19 [2]. By February 15, there were 23 cases, and then no new confirmed COVID-19 cases were reported until February 28 [3]. The group of cases up to February 15 represented the first wave of cases in Malaysia.

Subsequently increasing number of symptomatic individuals with presented to the healthcare facilities and were tested for the novel coronavirus. Those who tested positive were subsequently isolated and treated. A second wave of cases began on February 28, and since then the cumulative number of people affected by the coronavirus had risen to more than 1,000.

Containment measures at this stage were isolation and treatment of cases in dedicated COVID-19 hospitals coupled with active case detection among close contacts of the cases. In addition, individuals with history of travel to China or having close contacts with a confirmed COVID-19 case were classified as suspected cases and put under quarantine for two weeks. Other measures of containment included thermal and health screening of people at all international entry points and travel restriction advice to China.

By the first week of March 2020, the number of cases began to steadily rise. During this time, the Ministry of Health Malaysia received information from the Brunei counterpart about a man who was diagnosed with COVID-19 in that country. This individual claimed to have had participated in a religious mass gathering at a mosque in the outskirts of capital city Kuala Lumpur from 27 February to 1 March 2020. The gathering was attended by approximately 16,000 people from several neighboring countries [4].

This news raised the alarm in Malaysia. Active case detection among the participants of that mass gathering was immediately commenced, in an unprecedented exercise involving thousands of people. It was later noted that these participants had already spread the disease to their family members and close acquaintances [5]. As a result, the number of cases began to sharply rise.

At this stage, containment measures were enhanced. As it was no longer feasible to identify all infectious individuals and their contacts in the attempt to slow the spread of disease, the government applied community-wide containment measures. These interventions ranged from measures to encourage personal responsibility to identify disease and to increase social distancing among community members including cancellation of public gatherings. Despite these measures, the number of confirmed cases continued to rise. The government them went into mitigation phase.

In mitigation phase, the country was placed under country-wide 14-day quarantine from March 18 to March 31, under the Movement Control Order (MCO) [6]. The MCO included firstly general prohibition of mass movements and gatherings across the country including religious, sports, social and cultural activities. All premises must be closed except for supermarkets, public markets, convenience stores and convenience stores selling everyday necessities. Secondly, those who have just returned from overseas, they are required to undergo a health check and to do a quarantine (or self-quarantine) for 14 days; third, restrictions on the entry of all tourists and foreign visitors into the country; fourth, closure of all kindergartens, schools and pre-university institutions; fifth, the closure of all public and private higher education institutions; and sixth, the closure of all government and private premises except those involved in essential services (such as water, electricity, energy, telecommunications, postal, transportation and food supply).

During this phase, allocation of resources to the hospitals were enhanced: more intensive care unit beds were prepared, more human resources were deployed, and more personal protective equipment were acquired [7,8].

After having taken such unprecedented actions in controlling the epidemic, the forecast of the trend of this epidemic is crucial. The government needs to know if these actions had worked, and if so, to what extent. Forecast of the course of this epidemic is required in order to plan further on the actions that need to be taken.

One of the ways to forecast the trend of an epidemic is by using mathematical modelling, using either the deterministic or stochastic approaches. Deterministic model is based on the average characteristic of the population parameters under study, whereas stochastic model takes into account the randomness of elements of the population. Although stochastic model is more accurate in evaluating real-life epidemic propagation it is not entirely reproducible. Moreover, when the population is large enough these kinds of randomness neutralise each other and then a simpler deterministic model turns to be good enough to use. Simulation study based on deterministic compartmental model is most applied in epidemiology. Of the many deterministic models, the Susceptible-Exposed-Infectious-Removed (SEIR) models are often implemented when studying the spread of infectious diseases that possess significant incubation periods. In the case of COVID-19 several studies from China have also used this SEIR model [9,10, 11, 12].

Therefore, the aim of this paper is to forecast the progression of COVID-19 epidemic in Malaysia, before and after the Movement Control Order (MCO), using the SEIR model.

## Materials and methods

### Data

The data for this forecasting analysis was obtained from publicly available database and websites; mainly those of the Ministry of Health Malaysia and Department of Statistics Malaysia. Data included in this analysis are those from March 17, 2020 until March 27, 2020. This period was the first 10 days of the MCO exercise; and it was chosen because it coincided with the second wave of the epidemic in Malaysia. Two analyses were carried out, the first estimated the epidemic trend of COVID-19 for Malaysia before the MCO was implemented, while the second forecasted the trend after the implementation of MCO.

### SEIR Model

A deterministic SEIR model based on the clinical progression of the disease, epidemiological status of the individuals, and intervention measures was proposed. The mathematical model used was the Susceptible, Exposed, Infectious and Removed (SEIR) model, as shown in Fig. 1.

**Fig 1.**
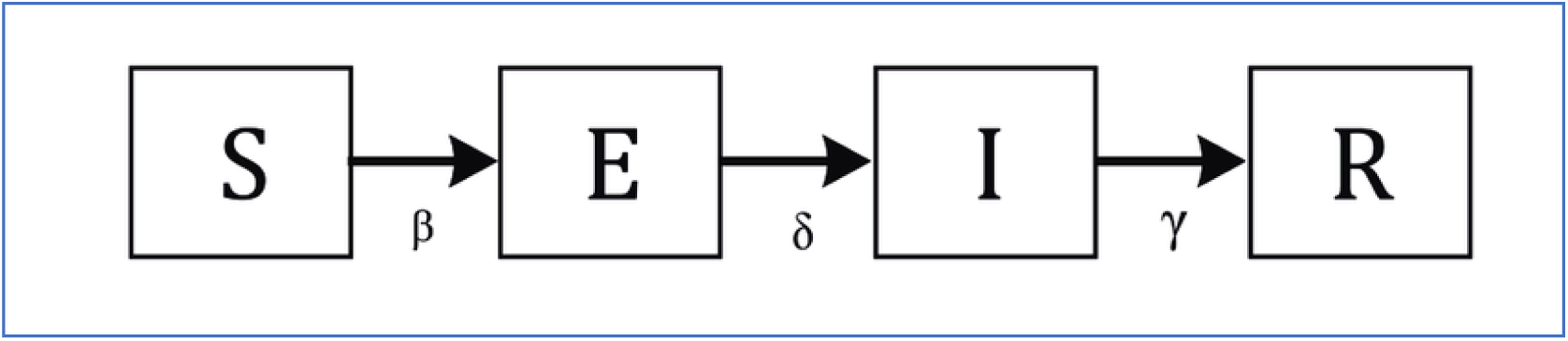
The Susceptible (S), Exposed (E), Infectious (I) and Removed (R) (SEIR) model.

The Susceptible, Exposed, Infectious and Removed are states in which an individual progresses in sequence. The infectious rate, β, controls the rate of spread which represents the probability of transmitting disease between a susceptible and an infectious individual. The incubation rate, *δ*, is the rate of latent individuals becoming infectious (average duration of incubation is 1/*δ*). Recovery rate, γ = 1/D, is determined by the average duration, D, of infection.

The quantity *S* denotes the number of individuals those are susceptible to the disease but not infected at time *t*. This is the total number of population who are at risk. The quantity *E* denotes the number of individuals those are exposed to the virus or infected but not yet infectious. This is the total number of population who came in contact with a disease person and carrying the infective agent. The quantity *I* denotes the number of infected individuals who can spread the disease through contact with susceptible. This is the number of exposed population developing sign and symptoms and infectious to others. The quantity *R* denotes the number of individuals those have successfully gained immunity from the disease and / or removed by death. This is the number of infected populations recovering from the disease and no longer infectious to others. The parameter *β* is transmission rate of disease from susceptible to exposed. Sigma (σ) refers to the Infection rate, while *γ* is the average durations infectiousness. The infectious diseases spread from an infected individual to other susceptible individuals in the surroundings. The governing equations describing the evolution and dynamics of SEIR model can be described by a set of ordinary differential equations as follows [13].

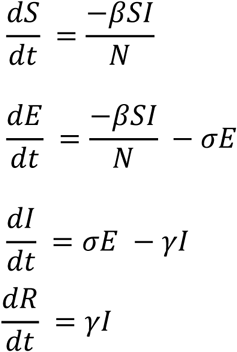

### Data analysis

Data included in this analysis are those from March 17, 2020 until March 27, 2020. It was chosen because it coincided with the second wave of the epidemic in Malaysia. The MCO also began at about the same time, which was on March 18.

In this model, the state susceptible (S) is the adult population aged 15 years and more, in Malaysia. This age range was chosen because so far reports have shown that COVID-19 has affected children and adolescents less than adults. Population in 2019 was approximately 32, 000,000 and of which, population in 2019 who were ≥ 15 years old was 76.7%. The exposed (E) is the number of people screened as of March 28 (35,516 people) minus the number of people screened as of March 17 (9,799 people). The infected (I) is the number of confirmed cases from March 17 until March 27, while the removed (R) is number of individuals those have successfully gained immunity from the disease and / or removed by death from March 17 until March 27. Table 1 summarizes the values and references of each of the states.

**Table 1.** Different parameters in the SEIR model and their values.

| Parameters | Value used for this analysis | Source |
| --- | --- | --- |
| Susceptible (S) | $S = 25,000,000$<br>S rate<br>$= 25,000,000/32,000,000$ | [14] |
|  | = 0.78125 |  |
| Exposed (E) | E = 25, 717<br>E rate<br>= 25, 717/32,000,000<br>= 0.0008 | [15, 16] |
| Infected (I) | I = 1608<br>I rate<br>= 1608/32,000,000<br>= 0.00005 | [15,17] |
| Recovered/died (R) | R= 216<br>R rate<br>= 216/32,000,000<br>= 0.000006 | [17] |

The main difference between the pre-MCO period and the MCO period is the reduced contact among individuals within the MCO period, compared to that in the before the MCO period. Other public health prevention and control measures remained the same.

In the context of the SEIR model, the MCO would affect the contact rates between the positive COVID-19 patients and the people around them, through the parameter Exposed (E)’s co-efficient which is beta (β). Beta value during the pre-MCO period used in this analysis was 6.47, based on the study in China [18]. The beta value of 6.47 was higher than other published estimates for example a value of 2.2 [19]. Such a high reproduction number was consistent with the opinion that the virus has gone through at least three to four generations of transmission in the period covered by their study [18,20] WHO, 2020). This stance was also held in this current study based on the findings by the Ministry of Health, that by the time the MCO was implemented there were at least 4 generations of infected individuals originating from the individuals who attended the mass gathering in late February 2020. Therefore, the first analysis using the SEIR model used the beta value of 6.47 to forecast the epidemic before the MCO in Malaysia was implemented.

We used the same dataset to forecast the epidemic trend *after* the implementation of the MCO. As mentioned, after the implementation of MCO the rate of contact between the infected and the exposed was anticipated to be lesser than the before the MCO period. The reduced rate of contact would result in a lower value for beta. A study estimated that the median daily reproduction number (*R*_t_) in Wuhan declined from 2·35 (95% CI 1·15–4·77) one week before travel restrictions were introduced on January 23, 2020, to 1·05 (0·41–2·39) one week after [21]. This was a 55% decline in the value of beta. In Malaysia, due to the limited movement among the population of Malaysia and the nationwide travel restrictions imposed through the MCO, we forecast that the epidemic after the MCO would also result in a reduction of the beta. Assuming that the reduction in beta was 55% from the without MCO, as in the abovementioned study [21] we set the beta value at 2.91, in the second analysis using the SEIR model.

For the coefficient of latency (σ) is 1/ period of latency. Because the latency period is yet not known for COVID-19, the value for incubation period was used. The incubation period was assumed to be Erlang distributed with mean 5·2 days (SD 3·7) (Li et al, 2020). The coefficient of migration rate (γ), is 1/ average recovery time. The average duration between the confirmation of diagnosis to date of removal (died/discharged) for Malaysia was 9.7 (± 5.3) days, based on sample of 58 patients from the start of the epidemic until 26.3.2020. However, patients may have had symptoms prior to presentation at the hospital or prior to the diagnosis confirmed. According to a study the median time from date of onset of symptoms to date of confirmation was 4.8 (± 3.0) days [22]. Therefore, the total average recovery time was the sum of 9.7 and 4.8 days which was equal to 14.5 days. Table 2 shows different coefficient in the SEIR model and their values.

**Table 2.** Different coefficient in the SEIR model and their values.

| Coefficients | Value used for this analysis | Source |
| --- | --- | --- |
| $\beta$ = coefficient of infection | $\beta = \beta_0 * k$<br>$\beta$ = coefficient of infection rate (R)<br>$\beta_0$ = probability of infection per exposure ($R_0$)<br>$k$ = frequency of exposure<br>$\beta^1 = 6.47$<br>$\beta^2 = 1.05$ | Tang et al, 2020<sup>1</sup><br>Kucharski et al, 2020<sup>2</sup> |
| $\sigma$ = coefficient of latency | $T_e$ = average latency<br>= 5.2 days<br>$\sigma = 1/ T_e$ | Li et al, 2020 |
| | $= 1/5.2$<br>$= 0.19$ | |
| $\gamma$ = coefficient of<br>migration rate | $\gamma = 1/ T_i$<br>$T_i$ = average recovery time<br>$T_i = 14.5$ days<br>$\gamma = 1/ T_i$<br>$= 0.07$ | Kraemer et al, 2020 |

In both analyses before the MCO and after MCO, it was assumed that the other parameters and their co-efficient would remain the same. The Susceptibility (S) rate depends on the population size with is massive, the Infection (I) rate would be the same it depends on the incubation period of the virus and Removal (R) rate would also remain the same as it depends on the recovery time. Because during the time of this analysis there was no introduction of any new drugs, the average recovery period would remain the same. Based on the available data and the above assumptions, an R software, R-3.6.3 with package “deSolve” was used to create the SEIR model [23].

## Results

At present there are two waves of the COVID-19 epidemic in Malaysia as shown in Fig. 2. The first wave was in early February, while the larger second wave began in late February.

**Fig. 2.**
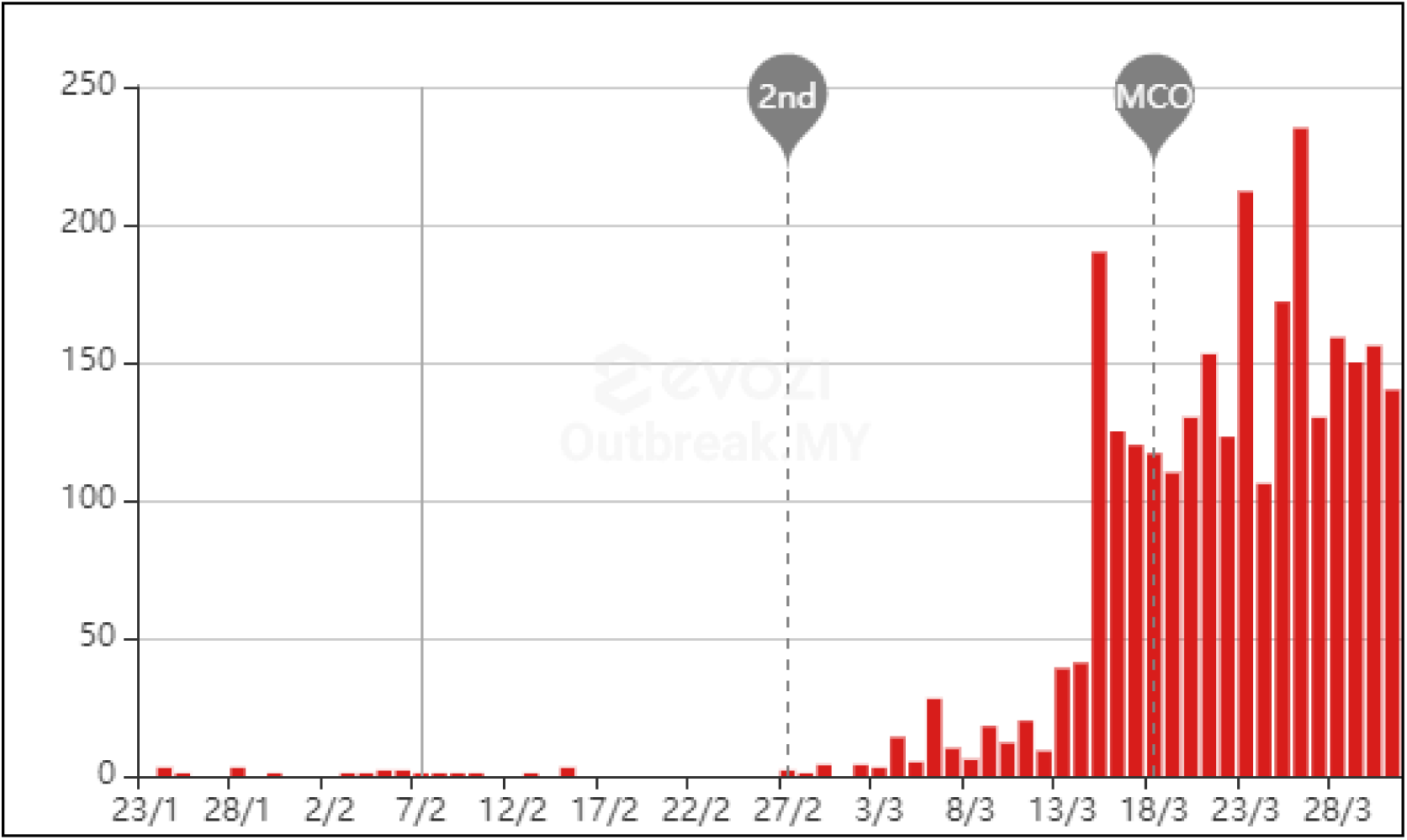
Distribution of confirmed new cases of COVID-19 in Malaysia. Source: [24]

Using the data and parameters as detailed in Table 1 in the SEIR model, the following graph was produced (Fig. 3). The graph shows that without the MCO, the number of new cases of infection would have peaked at day 18 after March 18 and would gradually subside and becomes plateaued at day 80 before finally ends by day 94 after March 18. At its peak, the infection could affect up to about 43% of the population.

**Fig. 3.**
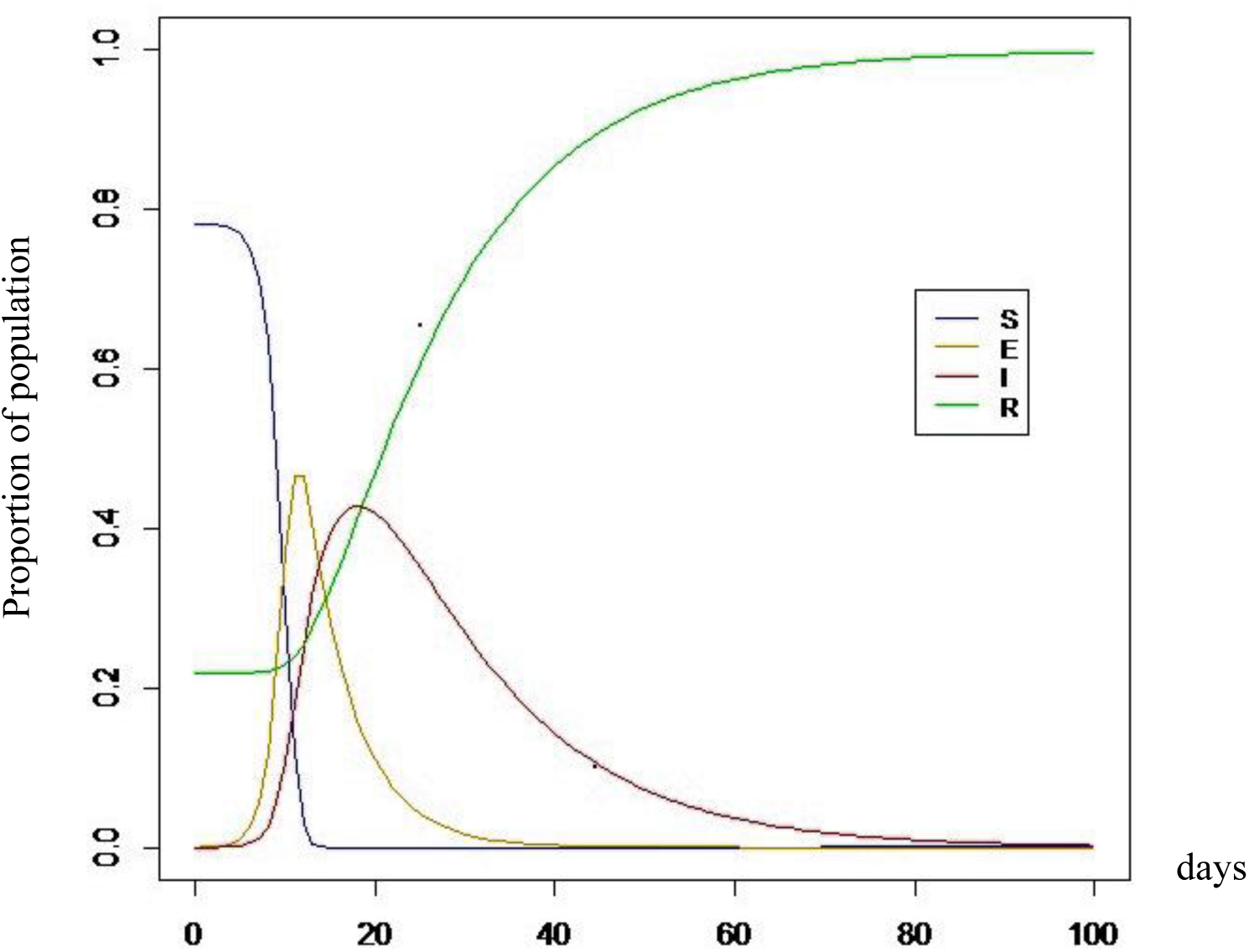
Forecast of the COVID-19 epidemic in Malaysia without MCO (beta = 6.47)

The above analysis was repeated using a smaller beta value to address the lesser contact rates among the susceptible population due to the implementation of the 14-day MCO. The result is shown in Fig. 4. The graph shows that with the implementation of MCO for 14 days, the number of new cases of infection would have peaked at about day 25 after March 31 and would gradually subside and becomes plateaued at day 90 after March 31 before finally ends by day 100 after March 31. At its peak, the infection could affect up to about 40% of the population.

**Fig. 4.**
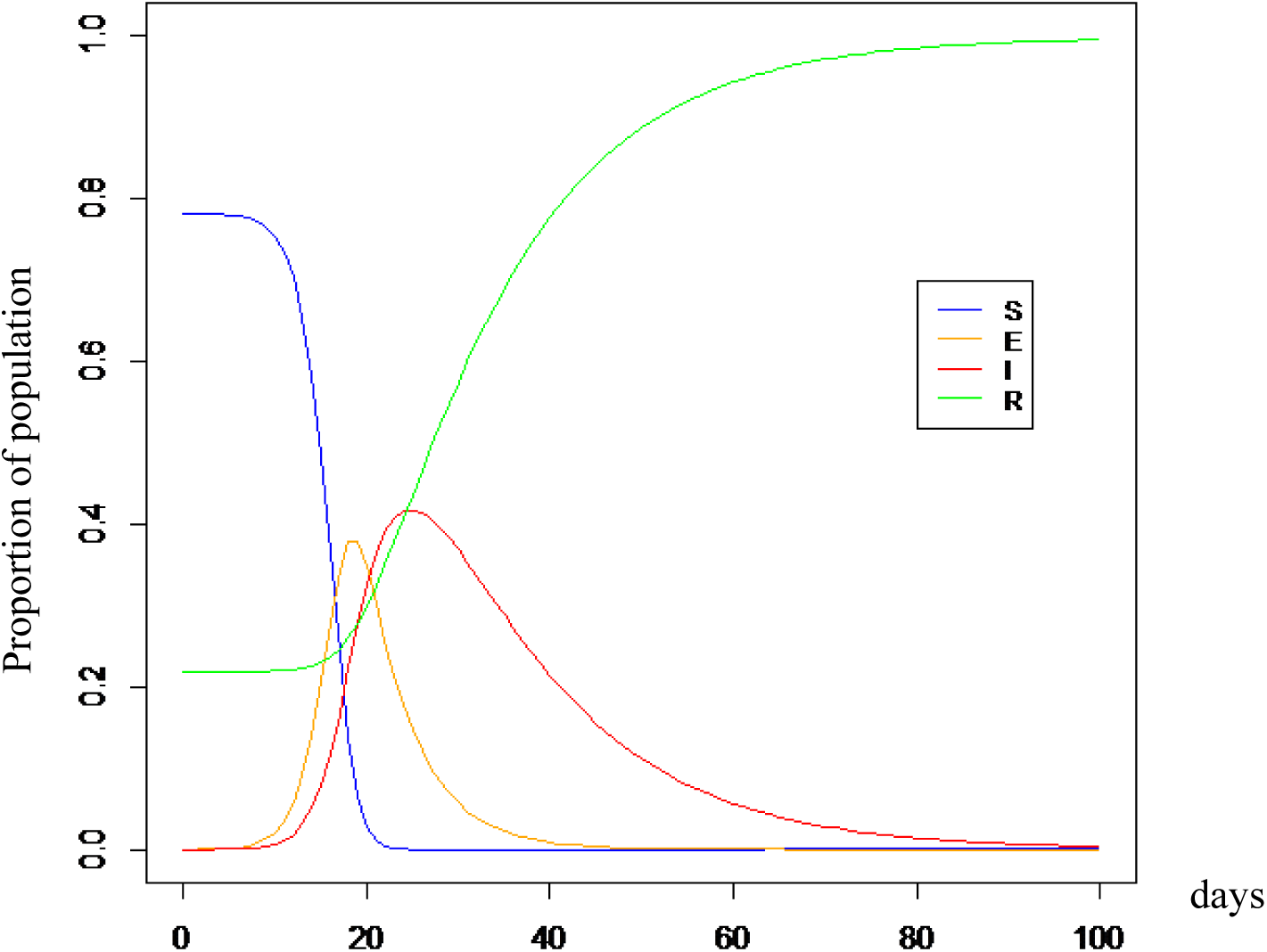
Forecast of the COVID-19 epidemic in Malaysia after 14-day MCO (beta = 2.91)

After the implementation of the14-day MCO, with a correspondingly lower beta value, we predict that there is a lesser peak in the number of people being infected but a slightly longer duration for the infection to subside. Summary of differences in parameters between pre-MCO and after MCO is shown in Table 3.

**Table 3.** Summary of differences in parameters between pre-MCO and after MCO.

| Parameters | Pre-MCO | After 14-day MCO |
| --- | --- | --- |
| Peak infection | Day 18 (9 April 2020) | Day 25 (25 April 2020) |
| Plateau | Day 80 (6 June 2020) | Day 90 (29 June 2020) |
| Subside | Day 94 (20 June 2020) | Day 100 (9 July 2020) |

## Discussion

The rationale of limiting movement of the population in an epidemic situation is to minimize the transmission of the infectious agent. Minimizing the transmission of the infectious agent would be favorable to the health system as it will reduce or avoid the surge of patients coming to health care facilities for treatment. The surge capacity of hospitals is often limited and the more than usual number of patients could result in substandard care due to limited disposable resources. Therefore, efforts like locking down a city or an entire province like what China did recently; or Movement Control Order (MCO) implemented in Malaysia, will to an extent, result in “flattening the epidemic curve”.

The COVID-19 curve is steep curve, in which the virus spreads exponentially (that is, case counts keep doubling at a consistent rate), and the total number of cases rise steeply to its peak within a few weeks. Infection curves like that of the COVID-19 with a steep rise also have a steep fall. A flatter curve, on the other hand, assumes the same number of people ultimately get infected, but over a longer period. A slower infection rate means a less stressed health care system. So, although the health authorities and governments alike are chanting the “let’s flatten the curve” mantra, one often neglected fact is that flattening the curve would also mean longer duration of the epidemic. As shown in our analysis, there should be reductions in the number of infected cases at the epidemic’s peak after the MCO effort, but the duration it takes for the epidemic to subside would be longer.

If the 14-day MCO in Malaysia was not implemented, based on the data analyzed and the assumptions used in the analysis, the peak of the infection period would be about 20 days after March 18, which was about the first week of April. At the peak period, the proportion of the population affected would be about 43% of susceptible individuals in the country. The duration of which new cases would continue to be recorded was predicted to be about 80 days from March 18 (approximately June 6, 2020). It is predicted that the epidemic would subside by about June 26, 2020.

With the implementation of the 14-day MCO in Malaysia beginning March 18 until March 31, the peak of the infection period would be about 30 days the end of the MCO (approximately April 30, 2020). At the peak period, the proportion of the population affected would be about 40% of susceptible individuals in the country. The duration of which new cases would continue to be recorded was predicted to be about 90 days from March 31 (approximately June 29, 2020). It is predicted that the epidemic would subside by about July 9, 2020. This forecast was again based on the data used and the assumptions that were used.

Whether or not the government chooses to prolong the duration of the MCO depends on whether it is best to suffer excruciating pain of an epidemic for a short period of time, or a milder but lingering pain of an epidemic for a longer period of time.

Longer duration of an epidemic could be detrimental to the economy. As an example, the lockdown in Hubei recently resulted in severe economic impact. Numerous firms evacuated their expat workers from the city and temporarily halted business activities, and among the industries that would be negatively impacted were retail, tourism, and hospitality sectors [25]. When China reopened its essential businesses on February 3, 2020, the markets dropped sharply in value [26]. The economic and labour crisis created by the Coronavirus Disease 2019 (COVID-19) pandemic could increase global unemployment by almost 25 million, according to a new assessment by the International Labour Organization (ILO). While at the national level, due to COVID-19 pandemic this year, Malaysia’s unemployment rate is expected to shoot up to 4% this year, from 3.3% in 2019. At the individual level, being unable to work and earn an income could lead the individual and his family into financial catastrophe. While at the government level, having to create economic stimulation packages to cushion the effect of the poor economy could cost a country, billions and even trillions of dollars. Even more challenging is that the stimulus package needs to be created in the circumstance when the government itself is not making any money.

In addition to the economic consequences, studies have shown that longer duration of movement restriction could result in negative psychological effects among all ages. A recent systematic review of literature on the psychological impact of quarantine in the general population showed that stressors included longer quarantine duration, infection fears, frustration, boredom, inadequate supplies, inadequate information, financial loss, and stigma; all of which could lead to depression and anxiety [27]. While for children and adolescents, stressors such as prolonged duration, fears of infection, frustration and boredom, inadequate information, lack of in-person contact with classmates, friends, and teachers, lack of personal space at home, and family financial loss can have more profound negative effects [27]. A study showed that the mean posttraumatic stress scores were four times higher in children who had been quarantined than in those who were not quarantined [28].

As for front-line health care workers, they too are also negatively affected. Scientific evidence on this is abundant. For example, a study in Japan found that during the H1N1 pandemic in 2009, workers with a high risk of infection felt more anxious and more exhausted [29]. Studies in China in 2003 during the SARS epidemic found that approximately 10% of hospital employees experienced high levels of posttraumatic stress (PTS) symptoms and that health care workers reported fatigue, poor sleep, worry about health, and fear of social contact, despite their confidence in infection-control measures; also have chronic stress and higher levels of depression and anxiety [30,31].

The limitations of this forecast lie mainly in the assumptions used and because the data used were from publicly available platforms. We used the data available on the day the data was announced. As there was a backlog of specimens to be analyzed of about 1,000 during the time of this analysis, we could not be certain whether the data on the new number of cases announced was entirely same-day data or it included data from specimens taken several days prior to the announcement. Apart from the number of new cases for each day, the nature of this data affected another parameter, namely the recovery period. For the recovery period, we considered the date of removal (discharged or death) and subtract that from the date of confirmation of diagnosis, instead of the actual dates of the start of illness to the day of discharge or death. This calculation may slightly under-estimate the duration of illness. Also, we did not consider the more frequent active case detection activities which have just been started during the production of this article. We also assumed that the laboratory capacity remains the same in the next few months, as when this analysis was conducted. Nonetheless, the results of this model could give an idea of the possible time span of the epidemic, until more detailed data is available. Additionally, the model could demonstrate that flattening the epidemic curve prolongs the duration of epidemic.

## Conclusion

After the implementation of the 14-day MCO from 18 March 2020 to March 31, it is forecasted that the epidemic in Malaysia will peak approximately in the end of April 2020 and will subside by about the first week of July 2020. Restricting movement of the population will reduce contact among individuals and hence would lessen the transmission and infection rates. This will “flatten the epidemiological curve” but will prolong the duration of the epidemic. Decision to extend the duration of the MCO should depend on the consideration of socioeconomic factors as well.

## Data Availability

All data referred to in the manuscript is made available with the manuscript.

## Supporting information

Files metadata

